# Retrieval-Augmented Claude Opus 4.7 and GPT-5.5 Surpass Human Performance on the Nuclear Cardiology Board Preparation Exam (and Claude Drafts a Paper About it)

**DOI:** 10.64898/2026.05.08.26352768

**Authors:** Aditya Killekar, Aakash Shanbhag, Robert JH Miller, Damini Dey, Paul B Kavanagh, Jamieson M. Bourque, Lawrence M. Phillips, Panithaya Chareonthaitawee, Piotr J. Slomka

## Abstract

**Background:** Previous studies evaluated large language model (LLM) performance on the American Society of Nuclear Cardiology (ASNC) Board Preparation Exam. Without domain-specific context, the best model (GPT-4o) achieved 63.1%, below the estimated 65% passing threshold and the 78% mean score of human fellows-in-training (FITs). Providing textbook context improved GPT-4o to 73.8% on text-only questions, but still fell short of human trainees. Whether next-generation LLMs with retrieval-augmented generation (RAG) can exceed this gap is unknown.

**Methods:** Claude Opus 4.7 and GPT-5.5 were administered all 168 questions (141 text-only, 27 image-based) from the 2023 ASNC Board Preparation Exam across 5 iterations each, using RAG with a nuclear cardiology textbook, companion atlas, and ASNC clinical guidelines. Claude used local FAISS-based semantic retrieval; GPT-5.5 used Azure’s cloud-hosted vector store. Performance was compared to prior LLM results and 13 human FITs.

**Results:** Across 5 iterations, Claude Opus 4.7 achieved a mean accuracy of 86.3% ± 1.4% (text 88.8%, image 73.3%). GPT-5.5 achieved 86.7% ± 2.2% (text 88.5%, image 77.0%) but refused a mean of 12.2 questions (7.3%) per iteration due to safety filters. Both models surpassed the human FIT mean (78.0%) and the estimated passing threshold. Compared to GPT-4o without context (63.1%), this represents a 23-percentage-point improvement in 18 months.

**Conclusion:** Next-generation LLMs with RAG now surpass average human trainee performance on nuclear cardiology board preparation questions, suggesting significant potential as educational tools and knowledge-reference aids in cardiovascular imaging.

**Condensed Abstract:** Across 5 iterations each, Claude Opus 4.7 and GPT-5.5 with retrieval-augmented generation achieved mean accuracies of 86.3% and 86.7% on the 2023 ASNC Board Preparation Exam (168 questions), both surpassing the mean human fellow-in-training score of 78%. GPT-5.5 refused a mean of 12.2 questions (7.3%) per iteration due to safety filters. These results represent a 23-percentage-point improvement over the best prior LLM without context (63.1%), demonstrating that RAG-enhanced LLMs have reached human-level proficiency in nuclear cardiology knowledge.

**Graphical Abstract:** 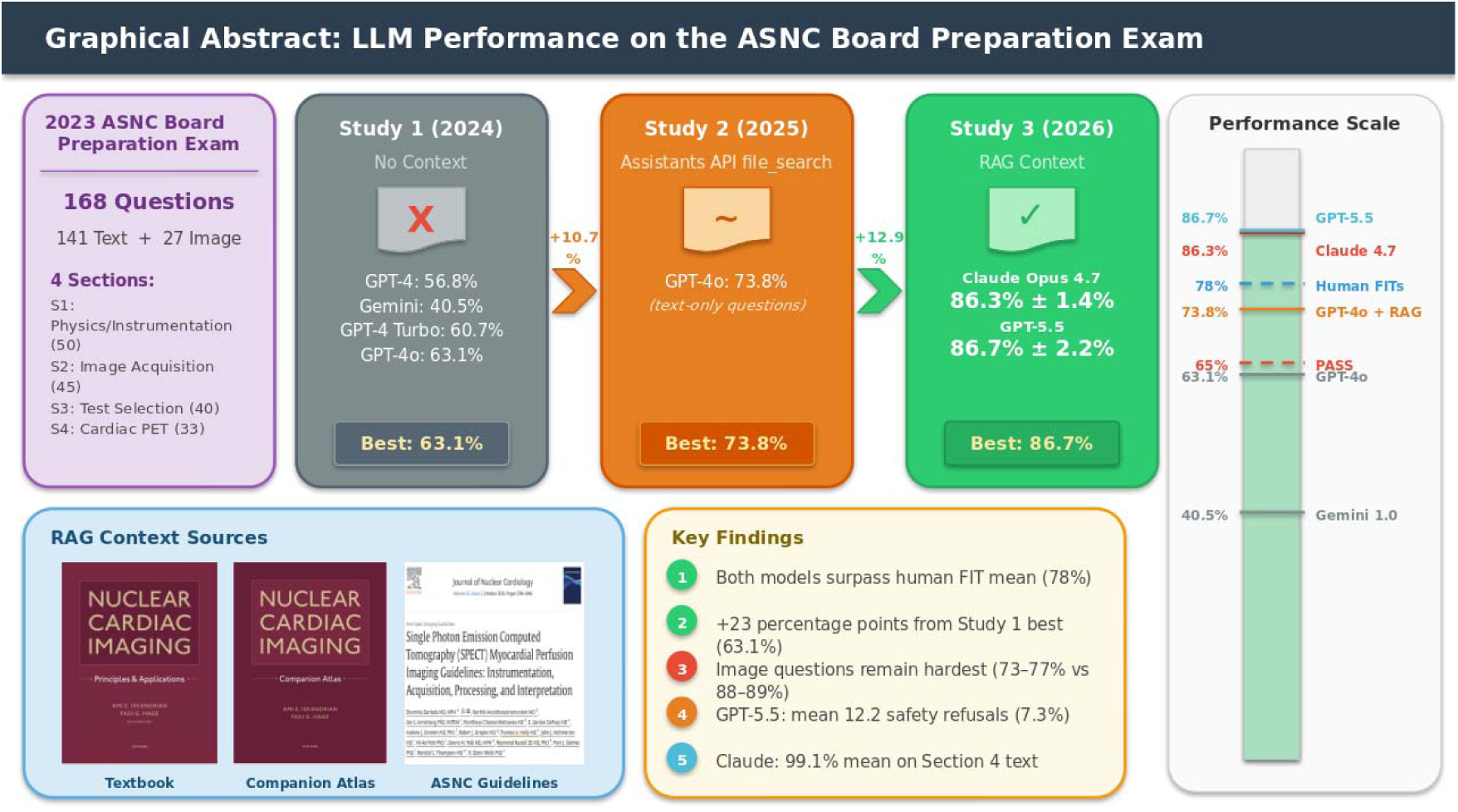

Overview of the three-study research arc evaluating LLM performance on the 2023 ASNC Board Preparation Exam. Study 1 (2024) tested four LLMs without context (best: GPT-4o, 63.1%). Study 2 (2025) added textbook context to GPT-4o (73.8%). Study 3 (2026, current) evaluated Claude Opus 4.7 and GPT-5.5 with retrieval-augmented generation across 5 iterations each (mean 86.3% and 86.7%, respectively), both surpassing the human fellow-in-training mean of 78%. Right panel shows the performance scale with key thresholds.

## INTRODUCTION

Large language models (LLMs) — artificial intelligence systems that use deep neural networks to learn from large quantities of textual information — have rapidly emerged as tools with potential applications across medicine, including medical education, clinical decision support, and diagnostic assistance (1, 2, 3). Their increasing scale, multimodal capabilities, and ability to process both text and images have prompted investigations into their performance on medical board examinations across multiple specialties (4–7). However, nuclear cardiology presents a uniquely challenging domain for LLMs due to its reliance on both specialized textual knowledge and nuanced interpretation of medical images including SPECT, PET, and multimodality imaging.

In our initial study (Study 1), we evaluated four LLMs — GPT-4, Google Gemini, GPT-4 Turbo, and GPT-4o (9) — on the 2023 ASNC Board Preparation Exam, which comprises 168 multiple-choice questions designed to reflect the content outline of the CBNC certification examination (8). Each model was tested 30 times to account for stochasticity. GPT-4o demonstrated the best overall performance with a median accuracy of 63.1% (95% CI 62.5–64.3%), significantly outperforming the other three models. However, this score fell below the estimated 65% passing threshold (10) and substantially below the mean score of 78.0% achieved by 13 human nuclear cardiology fellows-in-training (FITs) who completed the same examination.

In a subsequent research letter (Study 2), we investigated whether providing GPT-4o with domain-specific context — a nuclear cardiology textbook (*Nuclear Cardiac Imaging: Principles and Applications*), its companion atlas, and ASNC Clinical Practice Guidelines — would improve performance (11). When provided with these reference materials and tested at the optimal temperature setting (temperature = 0.01), GPT-4o achieved a median accuracy of 73.8% on text-only questions across 10 iterations per exam. This approach used the Azure OpenAI Assistants API with its built-in file search retrieval tool — an early form of retrieval-augmented generation, though with less user control over the retrieval pipeline than the explicit RAG implementations in the current study. This represented a meaningful improvement of approximately 10 percentage points over the no-context condition, but remained below the human FIT mean, and notably, this analysis was restricted to the 141 text-only questions.

Since our initial studies, the LLM landscape has evolved rapidly with the release of substantially more capable models. Retrieval-augmented generation (RAG) has emerged as a paradigm for grounding LLM responses in authoritative domain-specific knowledge, combining the reasoning capabilities of frontier models with curated reference materials retrieved at inference time (12, 13). The objective of the present study was to evaluate whether next-generation LLMs — Claude Opus 4.7 (Anthropic) and GPT-5.5 (OpenAI) — equipped with RAG pipelines accessing the same nuclear cardiology reference materials, could close or exceed the performance gap between LLMs and human trainees on the full 168-question ASNC Board Preparation Exam, including both text-only and image-based questions.

## METHODS

### Questions Dataset

The study utilized the same 168 multiple-choice questions from the 2023 ASNC Board Preparation Exam used in Studies 1 and 2. These questions were developed by expert Nuclear Cardiology faculty for ASNC’s Board Exam Preparation Course, constructed based on the CBNC exam content outline (14) and aligned with ASNC quality standards (15). ASNC granted permission to use these questions and provided the correct answers. The exam images are fully de-identified, contain no protected health information, and were uploaded through secured private accounts. Ethical/IRB approval was waived because there were no human subjects involved.

Questions were categorized into four sections according to ASNC guidelines: Section 1 — Physics, Instrumentation, Radionuclides, and Radiation Safety (n=50); Section 2 — Acquisition and Quality Control, Gated SPECT, Artifact Recognition, and MUGA (n=45); Section 3 — Test Selection, Stress and Nuclear Protocols Interpretation, Appropriate Use, and Risk Stratification (n=40); and Section 4 — Cardiac PET, Multimodality Imaging, Cardiac Amyloidosis, Cases with the Experts: PET and SPECT (n=33). Questions were further categorized by the presence or absence of images: 141 text-only questions and 27 image-based questions requiring interpretation of nuclear cardiology images.

### Reference Materials

The same domain-specific reference materials used in Study 2 were employed: (1) *Nuclear Cardiac Imaging: Principles and Applications* (textbook) (17), (2) its companion atlas, and (3) ASNC Clinical Practice Guidelines for interpretation of SPECT myocardial perfusion imaging (16) and related topics. These documents were pre-processed into 3-page PDF segments to create a corpus of retrievable chunks suitable for both retrieval pipelines.

### Retrieval-Augmented Generation Pipelines

Two distinct RAG implementations were used, one for each model (**Figure 1**):

**Figure 1.**
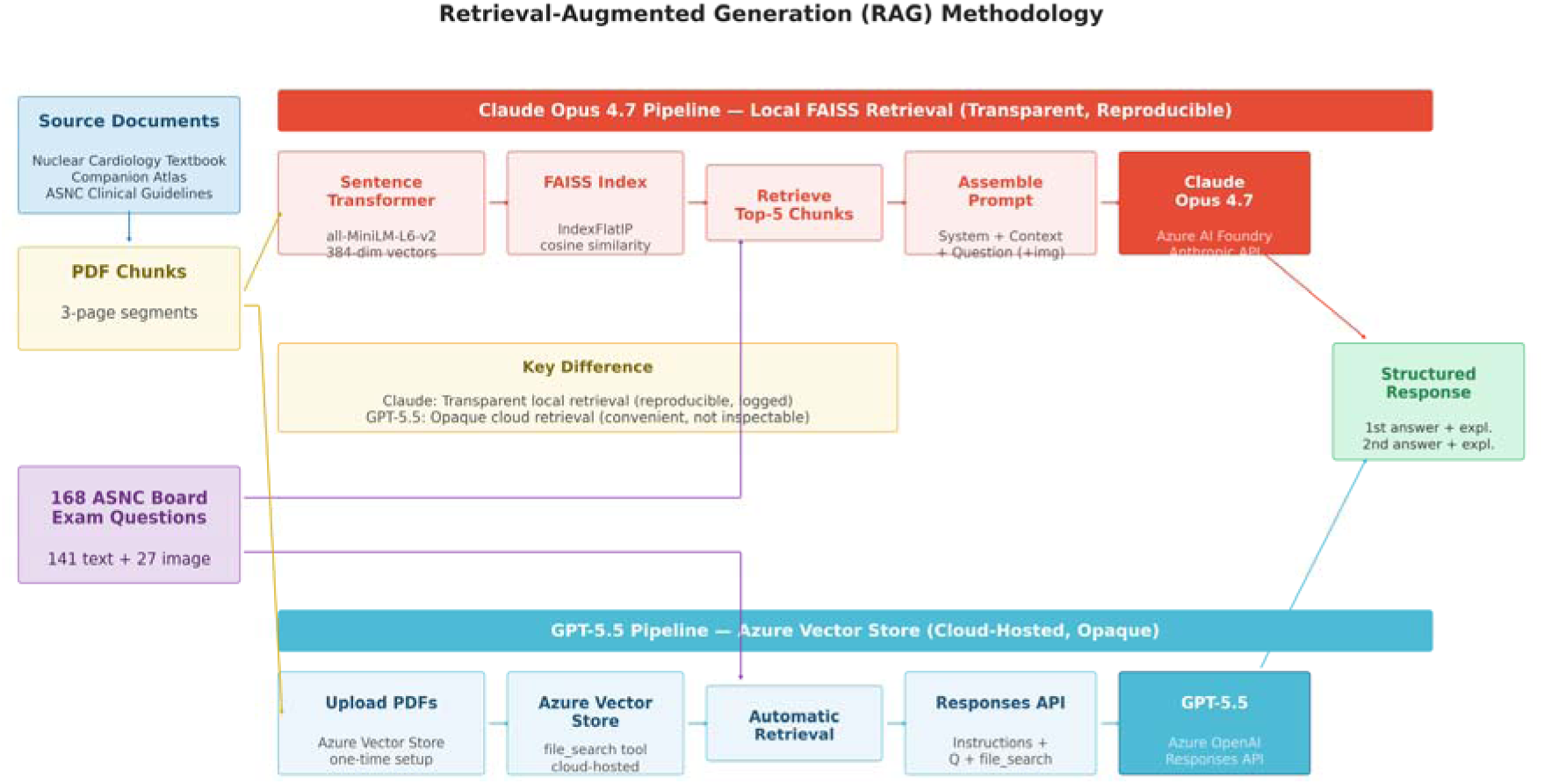
Retrieval-augmented generation methodology. Flow diagram showing the two parallel RAG pipelines: (top) Claude Opus 4.7 with local FAISS-based semantic retrieval using sentence-transformer embeddings and top-5 chunk retrieval; (bottom) GPT-5.5 with Azure cloud-hosted vector store and file_search tool. Both pipelines use the same source documents and exam questions.

#### Claude Opus 4.7 — Local FAISS Retrieval

The PDF chunks were embedded locally using the all-MiniLM-L6-v2 sentence-transformer model (18) (384-dimensional vectors) and indexed using a FAISS IndexFlatIP index (19), which performs exact inner-product search on L2-normalized vectors (equivalent to cosine similarity). For each exam question, the top 5 most semantically similar chunks were retrieved and prepended to the user message in the following format:

> “Use the following reference material to answer the question: [Context 1 — filename] [chunk text] [Context 2 — filename] [chunk text] … [up to 5 contexts] — [Exam question]”

This pipeline is fully transparent and reproducible: the embedding model, index type, retrieval count, and chunk sources are all specified and deterministic for a given query. An end-to-end example of context retrieval and prompt assembly for a sample question is shown in **Figure 2**.

**Figure 2.**
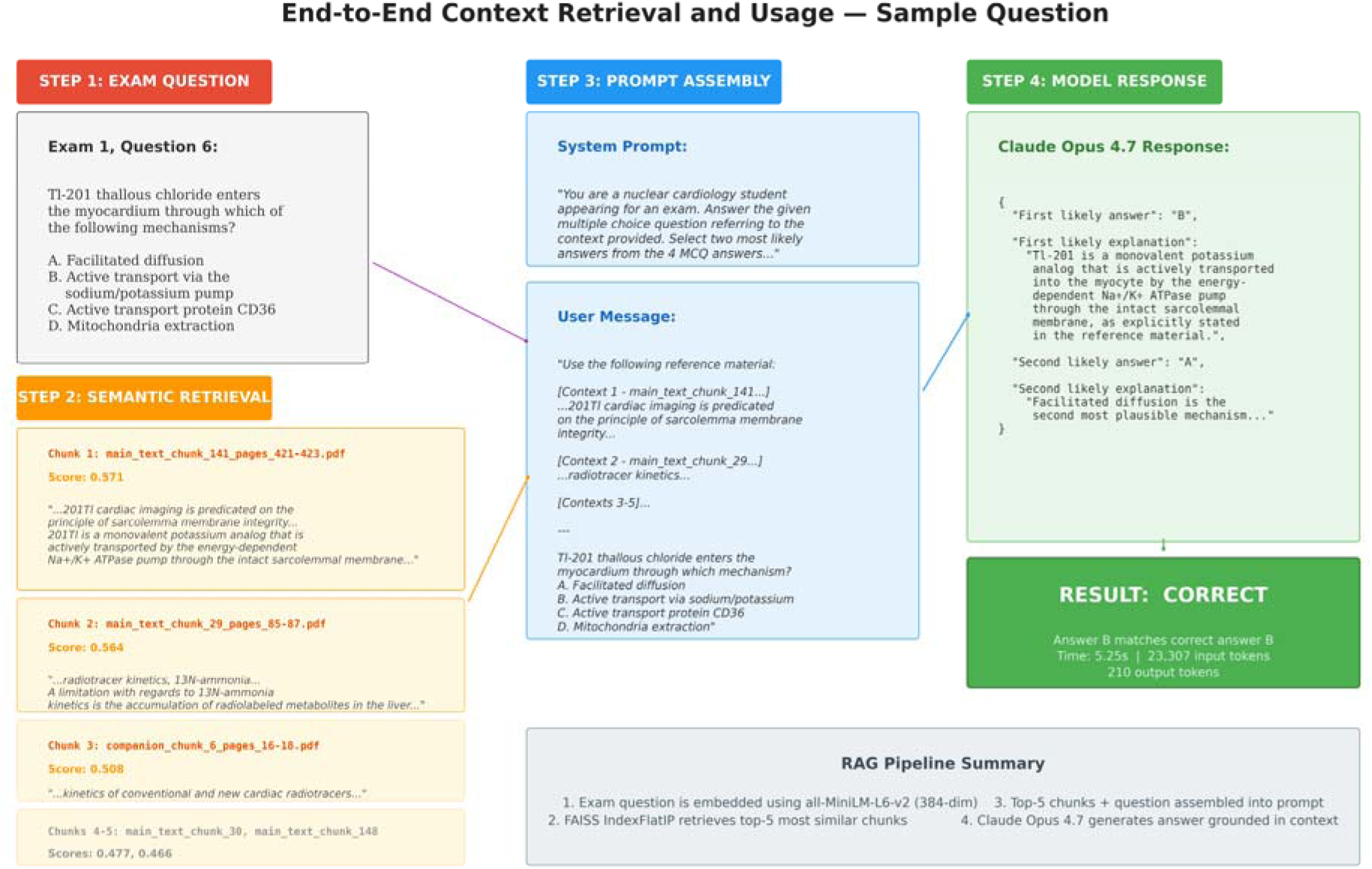
End-to-end context retrieval and usage for a sample question (Exam 1, Question 6: Tl-201 myocardial uptake mechanism). Shows the four steps: (1) exam question input, (2) semantic retrieval of the top-5 most relevant PDF chunks with similarity scores, (3) prompt assembly combining system instructions, retrieved context, and the question, and (4) model response citing the retrieved context to correctly identify active transport via the Na+/K+ ATPase pump

#### GPT-5.5 — Azure Vector Store Retrieval

The same PDF chunks were uploaded to an Azure OpenAI vector store using the vector_stores.file_batches.upload_and_poll API (API version 2025-04-01-preview). During inference, GPT-5.5 accessed these documents through the built-in file_search tool of the Azure OpenAI Responses API. Unlike the Claude pipeline, the retrieval mechanism — including the embedding model, similarity metric, and number of retrieved chunks — is managed by Azure’s cloud infrastructure and is not directly configurable or inspectable by the user.

### Model Configuration

**Claude Opus 4.7** (20) was accessed via the Azure AI Foundry Anthropic API using the AnthropicFoundry SDK. **GPT-5.5** (21) was accessed via the Azure OpenAI Responses API using the standard OpenAI SDK pointed at the Azure endpoint. The Assistants API does not support GPT-5.5; the Responses API was used as the supported approach.

Both models received a standardized system prompt:

> “You are a nuclear cardiology student appearing for an exam. Answer the given multiple-choice question referring to the context provided. Select two most likely answers from the 4 multiple choice answers for every question. The output should be strictly in the following format: {‘First likely answer’: ‘A/B/C/D’, ‘First likely explanation’: ‘…’, ‘Second likely answer’: ‘A/B/C/D’, ‘Second likely explanation’: ‘…’}”

For image-based questions, images were provided as base64-encoded data: Claude received images via the Anthropic vision API content blocks, and GPT-5.5 received images as input_image data URIs in the Responses API input payload. Both models had a maximum output token limit of 4,000 tokens. A 2-second delay was introduced between questions for API rate limiting.

### Exam Scoring

Each model was tested for 5 iterations across the full 168-question exam. The first most likely answer (A, B, C, or D) selected by each model was compared to the correct answer provided by ASNC. All questions were weighted equally. GPT-5.5 responses that were safety refusals (“I’m sorry, but I cannot assist with that request”) or empty responses were scored as incorrect. Scores are reported as means and standard deviations across iterations of the percentage of correctly answered questions overall, per exam section, and by question type (text-only vs. image-based).

### Comparison Groups

Results were compared descriptively to: (1) Study 1 models — GPT-4, Google Gemini, GPT-4 Turbo, and GPT-4o, each tested 30 times without domain-specific context; (2) Study 2 — GPT-4o tested 10 times per exam with textbook and guideline context at temperature 0.01; and (3) 13 human nuclear cardiology FITs from Study 1, who achieved a mean score of 78.0% (range 59.5–100.0%) with unlimited time and unrestricted access to external resources.

### Statistical Analysis

Descriptive statistics are presented as means, standard deviations, and ranges across 5 iterations per model. Comparisons across studies are descriptive given the differing experimental conditions (different models, different context approaches, different iteration counts). A question-level agreement analysis was performed for each iteration to characterize concordance and discordance between the two models on the 141 text-only questions; results are reported as ranges across iterations. All analyses were performed with Python 3.x using pandas, matplotlib, and seaborn.

### Independent Data Access and Analysis Statement

All authors had full access to the data and take responsibility for the integrity of the data and the accuracy of the data analysis.

## RESULTS

### Overall Performance

Across 5 iterations, Claude Opus 4.7 achieved a mean overall accuracy of 86.3% (SD 1.4%, range 85.1–88.1%). GPT-5.5 achieved a mean overall accuracy of 86.7% (SD 2.2%, range 83.3–88.7%). The two models performed at essentially identical levels (difference of 0.4 percentage points). Both models substantially exceeded the estimated 65% passing threshold for the CBNC examination in every iteration and surpassed the mean human FIT score of 78.0%. Compared to the best-performing model in Study 1 (GPT-4o, 63.1% without context), this represents mean improvements of 23.2 and 23.6 percentage points for Claude Opus 4.7 and GPT-5.5, respectively (**Figure 3**, **Table 2**).

**Figure 3.**
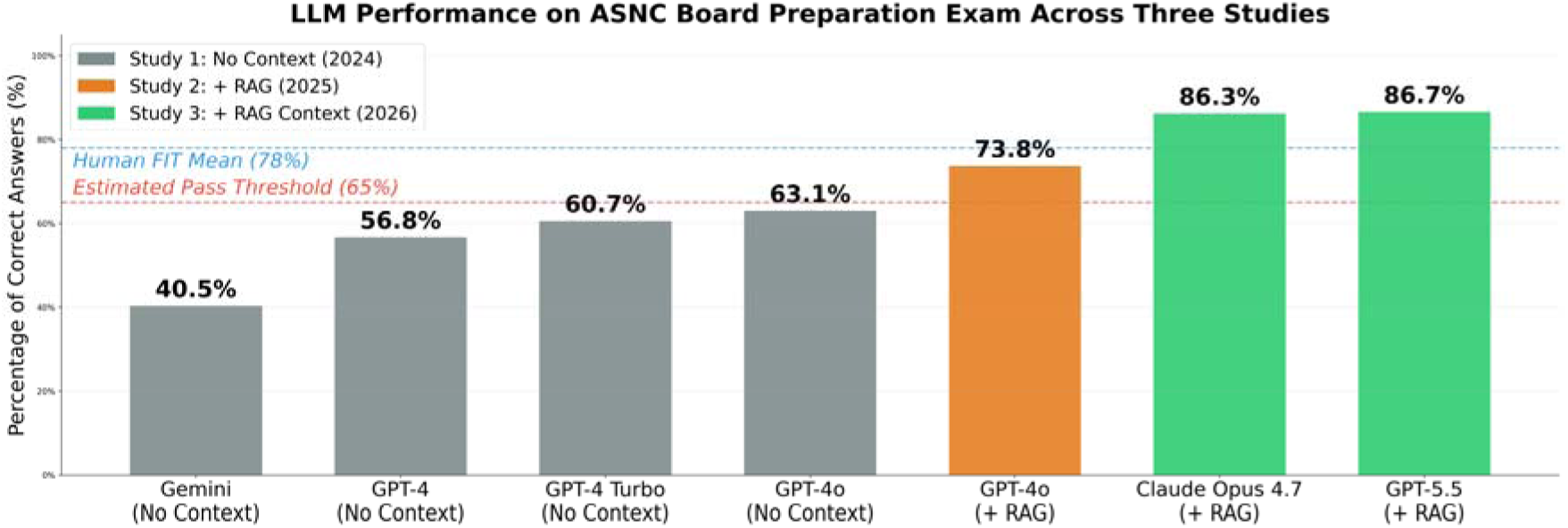
Longitudinal comparison of LLM performance across three studies. Bar chart showing overall accuracy (%) for all models evaluated across Studies 1–3. Horizontal dashed lines indicate the estimated 65% CBNC passing threshold (red) and the mean human FIT score of 78% (blue). Models are color-coded by study: gray (Study 1, no context), orange (Study 2, textbook context), green (Study 3, RAG context).

**Table 1.** Model Characteristics.

| Model | Developer | Study | Access Method | RAG Approach | Max Output Tokens |
| --- | --- | --- | --- | --- | --- |
| GPT-4 | OpenAI | 1 | Azure OpenAI API | None | 4,096 |
| Google Gemini | Google | 1 | Google AI API | None | 4,096 |
| GPT-4 Turbo | OpenAI | 1 | Azure OpenAI API | None | 4,096 |
| GPT-4o | OpenAI | 1, 2 | Azure OpenAI API / Assistants API | None (S1); Assistants API file_search (S2) | 4,096 |
| Claude Opus 4.7 | Anthropic | 3 | Azure AI Foundry Anthropic API | Local FAISS + sentence-transformers | 4,000 |
| GPT-5.5 | OpenAI | 3 | Azure OpenAI Responses API | Azure Vector Store (file_search) | 4,000 |

**Table 2.** Overall Performance Across All Studies.

| Model | Study | Context | N<br>Iterations | Overall<br>(%) | Text<br>(%) | Image<br>(%) |
| --- | --- | --- | --- | --- | --- | --- |
| Google Gemini | 1 | None | 30 | 40.5 | 44.7 | 22.2 |
| GPT-4 | 1 | None | 30 | 56.8 | 59.2 | 40.7 |
| GPT-4 Turbo | 1 | None | 30 | 60.7 | 62.4 | 44.4 |
| GPT-4o | 1 | None | 30 | 63.1 | 66.7 | 44.4 |
| GPT-4o | 2 | Textbook + Guidelines | 10 | 73.8* | 73.8 | N/A |
| GPT-5.5 | 3 | RAG (Azure Vector Store) | 5 | **86.7 ± 2.2** | **88.5 ± 2.7** | **77.0 ± 3.1** |
| Claude Opus 4.7 | 3 | RAG (FAISS Local) | 5 | **86.3 ± 1.4** | **88.8 ± 0.9** | **73.3 ± 4.1** |
| Human FITs | 1 | Unrestricted | 1 | 78.0 | 78.0 | 81.5 |

*Study 1 values are medians across 30 iterations. Study 2 values are means across 10 iterations per exam (text-only questions). Study 3 values are means ± SD across 5 iterations.*

### Performance by Exam Section

Both models performed consistently across all four exam sections, with mean accuracy ranging from 83.0% to 90.3% (**Table 3**, **Figure 4A**). Claude Opus 4.7 achieved the highest mean section-level accuracy on Section 4 (Cardiac PET, Multimodality Imaging; 90.3%, SD 1.4%), followed by Section 1 (Physics, Instrumentation; 88.0%, SD 2.0%), Section 2 (Acquisition and QC; 84.4%, SD 2.7%), and Section 3 (Test Selection, Stress Protocols; 83.0%, SD 2.1%). GPT-5.5 followed a similar pattern: Section 4 (89.7%, SD 1.7%), Section 1 (88.8%, SD 1.8%), Section 2 (84.4%, SD 3.5%), and Section 3 (84.0%, SD 6.0%).

**Figure 4.**
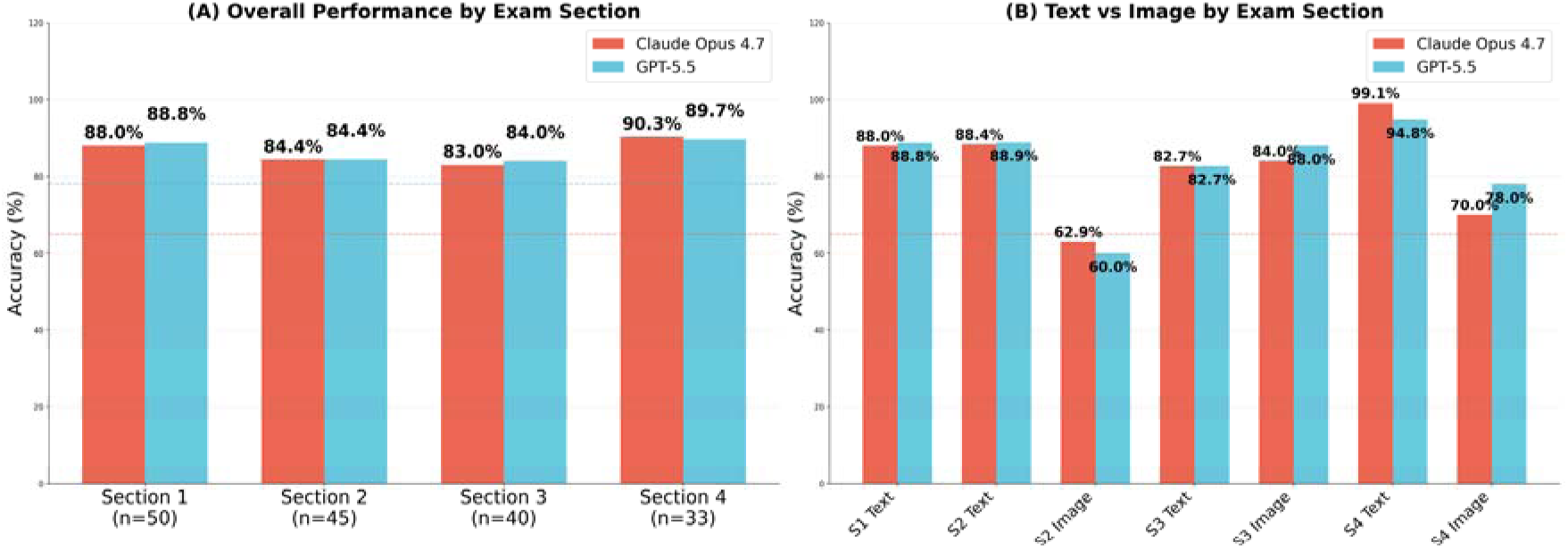
Per-exam section performance in Study 3. (A) Overall accuracy by exam section for Claude Opus 4.7 and GPT-5.5. (B) Text-only vs. image-based accuracy by exam section. Section 1 = Physics/Instrumentation (n=50); Section 2 = Acquisition/QC (n=45); Section 3 = Test Selection/Stress Protocols (n=40); Section 4 = Cardiac PET/Multimodality (n=33).

**Table 3.** Performance by Exam Section (Study 3)

| Exam Section | N Questions | Claude Opus 4.7 (%) | GPT-5.5 (%) |
| --- | --- | --- | --- |
| Section 1: Physics, Instrumentation | 50 | 88.0% ± 2.0% | 88.8% ± 1.8% |
| Section 2: Acquisition, QC, SPECT | 45 | 84.4% ± 2.7% | 84.4% ± 3.5% |
| Section 3: Test Selection, Stress | 40 | 83.0% ± 2.1% | 84.0% ± 6.0% |
| Section 4: Cardiac PET, Multimodality | 33 | 90.3% ± 1.4% | 89.7% ± 1.7% |
| **Total** | **168** | **86.3% ± 1.4%** | **86.7% ± 2.2%** |

*Values are means ± SD across 5 iterations.*

Both models demonstrated their strongest performance on Section 4, which covers cardiac PET and multimodality imaging. The lowest performance for both models was on Section 3, which requires integration of clinical decision-making with nuclear cardiology protocols.

### Text-Only vs. Image-Based Questions

On the 141 text-only questions, Claude Opus 4.7 achieved a mean accuracy of 88.8% (SD 0.9%) and GPT-5.5 achieved 88.5% (SD 2.7%). On the 27 image-based questions, Claude Opus 4.7 achieved a mean of 73.3% (SD 4.1%) and GPT-5.5 achieved 77.0% (SD 3.1%) (**Table 4**, **Figure 5**).

**Figure 5.**
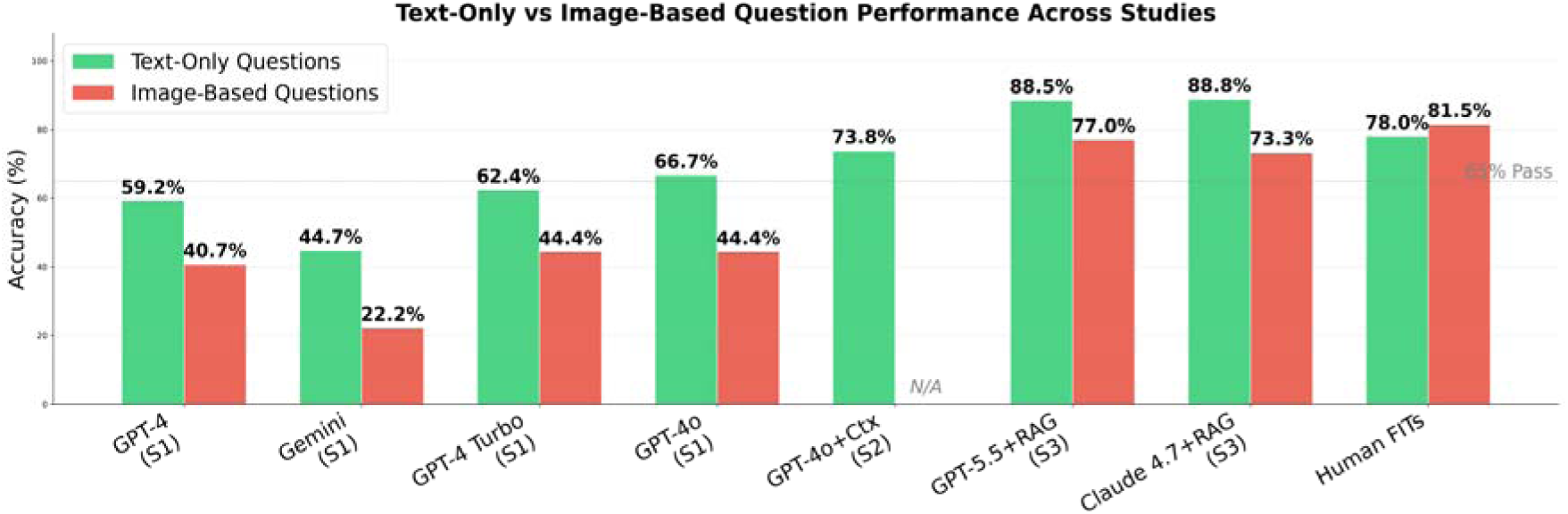
Text-only vs. image-based question performance across all studies and human FITs. Green bars represent text-only question accuracy; red bars represent image-based question accuracy. N/A indicates Study 2 did not test image-based questions.

**Table 4.** Text-Only vs. Image-Based Performance.

| Question Type | N | Claude Opus 4.7 | GPT-5.5 | GPT-4o + Context (S2) | GPT-4o No Context (S1) | Human FITs |
| --- | --- | --- | --- | --- | --- | --- |
| Text-only | 141 | 88.8% ± 0.9% | 88.5% ± 2.7% | 73.8% | 66.7% | 78.0% |
| Image-based | 27 | 73.3% ± 4.1% | 77.0% ± 3.1% | N/A | 44.4% | 81.5% |
| **Overall** | **168** | **86.3% ± 1.4%** | **86.7% ± 2.2%** | **73.8%** | **63.1%** | **78.0%** |

*Study 3 values are means ± SD across 5 iterations.*

The mean text-image performance gap was 15.5 percentage points for Claude Opus 4.7 and 11.5 percentage points for GPT-5.5. This pattern is consistent with Study 1, where all four models showed lower accuracy on image-based questions compared to text-only questions.

Notably, Claude Opus 4.7 achieved a mean of 99.1% on Section 4 text-only questions, with 4 of 5 iterations scoring a perfect 100% (23/23). GPT-5.5 achieved a mean of 94.8% on the same subset (**Figure 4B**).

### Model Agreement and Response Comparison

A question-level agreement analysis on the 141 text-only questions across 5 iterations revealed substantial concordance between the two models. Both models answered correctly on 78.0–83.0% of questions per iteration, and both answered incorrectly on 1.4–5.7%. Claude Opus 4.7 alone answered correctly on 5.0–9.9% of questions per iteration, and GPT-5.5 alone answered correctly on 5.7–9.9%.

When both models answered correctly, they typically provided concordant reasoning grounded in the retrieved context. For example, on Exam 1 Question 6 — “Tl-201 thallous chloride enters the myocardium through which mechanism?” — both models correctly identified active transport via the Na+/K+ ATPase pump (answer B), citing the retrieved textbook content on sarcolemmal membrane integrity (**Supplementary Figure S1A**).

When models disagreed, the errors typically reflected nuanced distinctions. On Exam 1 Question 26 — regarding how energy resolution is commonly reported — Claude Opus 4.7 incorrectly selected FWHM (answer B, confusing spatial resolution with energy resolution), while GPT-5.5 correctly identified percent FWHM (answer C) (**Supplementary Figure S1B**). In cases where both models erred, they frequently converged on the same incorrect answer, suggesting shared gaps in the retrieved context or reasoning patterns (**Supplementary Figure S1C**).

### GPT-5.5 Safety Refusals

Across 5 iterations, GPT-5.5 refused a mean of 12.2 of 168 questions per iteration (7.3%, SD 3.1, range 9–17), typically responding with “I’m sorry, but I cannot assist with that request.” All refusals were scored as incorrect. Claude Opus 4.7 had zero refusals across all 5 iterations (840 total responses) (**Supplementary Figure S2**).

In the first iteration, 13 refusals spanned all four exam sections: Section 1 (4 refusals), Section 2 (2), Section 3 (4), and Section 4 (3). Analysis of these refused questions revealed several thematic categories: radiopharmaceutical preparation and handling (3 questions — RBC labeling kit procedures, USP sterile preparation standards, and radiopharmaceutical dosing calculations), radiation safety including pregnancy-related dose limits (1 question), clinical patient management scenarios involving stress testing and imaging protocols (5 questions), pharmacology of stress agents (1 question — adenosine receptor subtypes), and nuclear imaging technical procedures (3 questions — thallium gating, PET interpretation, and cardiac viability assessment). No single category dominated, suggesting that the safety classifier is broadly sensitive to medical content involving radiopharmaceuticals, radiation, and clinical management rather than targeting a specific topic.

Had GPT-5.5 not refused any questions, its refusal-adjusted accuracy on answered questions would be approximately 93.4%, substantially higher than its observed mean of 86.7%. The refusal behavior thus represents a significant practical limitation, particularly given that the refused questions concerned legitimate nuclear cardiology examination content.

### Longitudinal Comparison Across Studies

**Figure 3** presents the longitudinal trajectory of LLM performance across all three studies. The progression shows a clear upward trend: from Gemini’s 40.5% (Study 1) through GPT-4o’s 63.1% (Study 1, best without context), to GPT-4o’s 73.8% with context (Study 2), and finally to mean accuracies of 86.3% (Claude Opus 4.7) and 86.7% (GPT-5.5) with RAG (Study 3). The overall improvement from the best no-context model to the best mean RAG-enhanced performance spans 23.6 percentage points in approximately 18 months.

### Comparison with Human Performance

The 13 human FITs in Study 1 achieved overall scores ranging from 59.5% to 100.0%, with a mean of 78.0% (131/168) (**Table 5**). Based on mean accuracies, GPT-5.5 (86.7%) would rank 5th and Claude Opus 4.7 (86.3%) would rank 6th (tied with FIT 12) among the combined 15 participants (13 FITs + 2 LLMs), exceeding 9 and 8 of the 13 human test-takers, respectively. On text-only questions, Claude Opus 4.7 at 88.8% and GPT-5.5 at 88.5% substantially exceeded the human FIT text-only mean of 78.0% (110/141). Notably, the human FIT image-based mean was 81.5% (22/27), higher than both models’ mean image accuracy (Claude 73.3%, GPT-5.5 77.0%), indicating that human trainees retain an advantage in image interpretation despite lower overall scores.

**Table 5.** Comparison with Human Fellows-in-Training.

| Rank | Participant | Overall (n=168) | Text (n=141) | Image (n=27) |
| --- | --- | --- | --- | --- |
| 1 | FIT 5 | 168 (100.0%) | 141 (100.0%) | 27 (100.0%) |
| 2 | FIT 10 | 157 (93.5%) | 132 (93.6%) | 25 (92.6%) |
| 3 | FIT 1 | 154 (91.7%) | 129 (91.5%) | 25 (92.6%) |
| 4 | FIT 11 | 151 (89.9%) | 126 (89.4%) | 25 (92.6%) |
| 5 | **GPT-5.5** | **86.7% ± 2.2%** | **88.5% ± 2.7%** | **77.0% ± 3.1%** |
| 6 | FIT 12 | 145 (86.3%) | 120 (85.1%) | 25 (92.6%) |
| 6 | **Claude Opus 4.7** | **86.3% ± 1.4%** | **88.8% ± 0.9%** | **73.3% ± 4.1%** |
| 7 | FIT 13 | 134 (79.8%) | 112 (79.4%) | 22 (81.5%) |
| 8 | FIT 6 | 131 (78.0%) | 109 (77.3%) | 22 (81.5%) |
| 9 | FIT 7 | 126 (75.0%) | 108 (76.6%) | 18 (66.7%) |
| 10 | FIT 3 | 124 (73.8%) | 106 (75.2%) | 18 (66.7%) |
| 11 | FIT 9 | 113 (67.3%) | 98 (69.5%) | 15 (55.6%) |
| 12 | FIT 2 | 107 (63.7%) | 87 (61.7%) | 20 (74.1%) |
| 13 | FIT 4 | 101 (60.1%) | 82 (58.2%) | 19 (70.4%) |
| 14 | FIT 8 | 100 (59.5%) | 77 (54.6%) | 23 (85.2%) |
|  | **FIT Mean** | **131 (78.0%)** | **110 (78.0%)** | **22 (81.5%)** |

## DISCUSSION

This study demonstrates for the first time that LLMs equipped with retrieval-augmented generation surpass the average performance of human nuclear cardiology fellows-in-training on the ASNC Board Preparation Exam. Across 5 iterations, Claude Opus 4.7 achieved a mean accuracy of 86.3% (SD 1.4%) and GPT-5.5 achieved 86.7% (SD 2.2%), representing a dramatic improvement from the 63.1% achieved by the best LLM just 18 months prior. This milestone — crossing the human trainee performance threshold — has significant implications for the potential role of LLMs as educational and knowledge-reference tools in nuclear cardiology.

### The RAG Paradigm Shift

The improvement observed across our three studies is attributable to two factors: more capable base models and retrieval-augmented generation grounding responses in authoritative domain texts. Study 2 demonstrated that providing context via the Azure Assistants API file_search tool boosted GPT-4o by approximately 10 percentage points, and the current study shows that next-generation models with RAG add another 13 percentage points. Disentangling the relative contributions of model advancement versus RAG improvement is not possible with the current study design — a controlled experiment using the same model with and without context would be needed.

The two RAG implementations used in this study — Claude’s transparent local FAISS pipeline with known embedding models and retrieval counts, and GPT-5.5’s opaque Azure cloud-hosted vector store — achieved comparable mean results (86.3% vs. 86.7%), with overlapping ranges across 5 iterations. This suggests that the RAG approach is robust across different implementations. The local FAISS pipeline offers reproducibility and transparency advantages (every retrieved chunk and similarity score is logged), while the cloud pipeline offers convenience. For research applications, the transparent approach is preferable; for clinical deployment, the trade-offs between transparency and ease of integration merit consideration.

### The Persistent Image Gap

Despite substantial improvements on text-based questions, image-based questions remain the most challenging category. The models achieved mean image accuracy of 73.3% (Claude) and 77.0% (GPT-5.5), which is 15.5 and 11.5 percentage points below their respective mean text-only accuracy. This gap is consistent with Study 1, where all four models performed worse on image-based questions. The image questions require interpretation of SPECT perfusion images, PET scans, polar maps, phase histograms, and multimodality imaging — tasks that demand spatial reasoning and pattern recognition beyond textual knowledge retrieval.

Notably, the mean image accuracy (73.3–77.0%) is substantially higher than in Study 1, where GPT-4o achieved only 44.4% on image questions. This improvement may reflect advances in the vision capabilities of newer models, though comparative studies of multimodal LLMs against radiologists (22) and systematic evaluations on medical visual question answering (23) suggest persistent limitations in AI image interpretation. Closing the remaining gap to text-level performance will likely require advances in medical vision models or domain-specific image fine-tuning, as models’ capabilities on image interpretation appear to stem primarily from general pre-training rather than targeted medical imaging training.

### GPT-5.5 Safety Refusals

A notable practical finding is GPT-5.5’s refusal to answer a mean of 12.2 of 168 questions per iteration (7.3%), typically with the message “I’m sorry, but I cannot assist with that request.” These refusals occurred across all four exam sections and affected both text and image questions. In contrast, Claude Opus 4.7 answered every question without refusal across all 5 iterations.

This behavior likely reflects overly conservative safety classifiers in GPT-5.5 that flag legitimate medical examination content as potentially harmful. The refused questions spanned radiopharmaceutical preparation (e.g., RBC labeling procedures, USP sterile preparation standards), radiation safety (pregnant technologist dose limits), clinical patient management (stress testing protocols, post-revascularization evaluation), and pharmacology (adenosine receptor mechanisms), consistent with the broader phenomenon of exaggerated safety behaviors documented in large language models (24). While safety guardrails are essential, their current calibration creates a real-world barrier to deploying GPT-5.5 in medical education contexts. A model that refuses to answer nearly 8% of legitimate board examination questions has a significant operational limitation that impacts both its utility and its observed accuracy.

### Clinical and Educational Implications

LLMs surpassing average trainee performance suggests several potential applications: (a) board examination preparation aids that can explain nuclear cardiology concepts with reference to authoritative sources, (b) knowledge-reference tools that may inform protocol selection and image interpretation in the nuclear cardiology reading room, (c) educational modules where LLM-generated explanations supplement traditional teaching, and (d) quality assurance tools for educational material development — their ability to answer questions correctly when provided with specific reference materials can help educators verify that training resources are sufficiently detailed and on-target for the topics being assessed.

However, examination performance does not equate to clinical competence. Real-world nuclear cardiology requires integrating patient-specific context, managing diagnostic uncertainty, communicating findings effectively, and exercising clinical judgment — capabilities that cannot be assessed through multiple-choice questions alone. We maintain that LLMs are best positioned as supplementary knowledge tools rather than replacements for physician expertise, consistent with our conclusions in Studies 1 and 2.

### Comparison with Other Specialties

Our findings align with a broader trend of LLMs achieving or surpassing human-level performance on medical board examinations. Studies have demonstrated strong LLM performance on the USMLE (4, 25), radiology board exams (6, 26), ophthalmology board questions (27), orthopaedic surgery certification exams (28), and cardiac imaging questions (29), with similar evaluations on Japanese board examinations in emergency medicine (30), otolaryngology (31), and radiology (32). Expert-level medical question answering has been achieved through domain-specific approaches including Med-PaLM 2 (33), and multimodal medical AI models such as Google’s Gemini continue to advance across clinical benchmarks (34). The nuclear cardiology domain is distinctive for its dual requirement of text-based medical knowledge and medical image interpretation, making the achievement of 86–87% mean accuracy particularly noteworthy in a field where multimodal reasoning is essential.

### Limitations

This study has several limitations:

1. **Limited iteration count.** The current study used 5 iterations per model, fewer than Study 1 (30 iterations) and comparable to Study 2 (10 iterations per exam). While 5 iterations allow calculation of means, standard deviations, and ranges, they provide limited statistical power for formal hypothesis testing or narrow confidence intervals. Notably, Claude Opus 4.7’s local FAISS retrieval pipeline is deterministic — given the same embeddings and query, the same chunks are retrieved every time — so variability in Claude’s responses stems only from model sampling, not retrieval differences. GPT-5.5’s cloud-hosted vector store introduces additional stochasticity in retrieval that is not user-controllable. Future work should include additional iterations repeated across multiple days to assess day-to-day performance drift.
2. **Potential data contamination.** The same 168 questions have been used across all three studies. Newer models trained on more recent data may have been exposed to these questions or similar content during pre-training, potentially inflating performance estimates. However, the questions were only available through ASNC’s Board Exam Preparation Course and are not publicly available online.
3. **Differing RAG implementations.** The two models used different retrieval mechanisms (local FAISS vs. cloud vector store), preventing a pure model-to-model comparison. Differences in retrieval quality, chunk selection, and context assembly may contribute to performance differences beyond the models’ intrinsic capabilities.
4. **ASNC exam as proxy.** The ASNC Board Preparation Exam, while aligned with the CBNC content outline, does not perfectly replicate the actual certification exam. The difficulty level, question format (no video-based questions), and content coverage may differ.
5. **Uncontrolled human comparison.** The 13 human FITs had unlimited time and unrestricted access to external resources, whereas the LLMs were provided only the curated context documents. This makes the comparison directionally informative but not strictly equivalent.
6. **Temperature and sampling parameters.** Default temperature settings were used for both models in the current study. Study 2 demonstrated that temperature significantly affects GPT-4o performance; the current results may not represent optimal performance for either model.
7. **Limited model diversity.** Only two next-generation models were tested. Other frontier models — including Google Gemini 2.0, Meta Llama, and domain-specific medical LLMs — were not evaluated and may show different performance profiles.
8. **Refusal impact on GPT-5.5.** The mean 12.2 safety refusals per iteration disproportionately affect GPT-5.5’s observed accuracy. If refused questions were excluded from the denominator, GPT-5.5’s accuracy on answered questions would be approximately 93.4%, substantially higher than its observed mean of 86.7%.

### New Knowledge Gain

#### What is new?

This is the first study to demonstrate that LLMs with retrieval-augmented generation surpass the average performance of human nuclear cardiology fellows-in-training on the ASNC Board Preparation Exam. - Across 5 iterations, Claude Opus 4.7 achieved a mean of 86.3% (SD 1.4%) and GPT-5.5 achieved 86.7% (SD 2.2%), representing a 23-percentage-point improvement over the best prior LLM without context in just 18 months. - Mean image accuracy was 73.3% (Claude) and 77.0% (GPT-5.5), demonstrating that medical image interpretation remains a persistent challenge regardless of the underlying model.

#### What are the clinical implications?

RAG-enhanced LLMs demonstrate sufficient nuclear cardiology knowledge to serve as educational aids for board preparation and as knowledge-reference tools. - Safety refusal behavior in GPT-5.5 (mean 7.3% of questions per iteration) highlights a practical barrier to deploying LLMs in medical education contexts that must be addressed by model developers. - The persistent gap between text and image question performance indicates that LLMs are not yet ready to replace human expertise in diagnostic image interpretation.

## Data Availability

N/A

## ACKNOWLEDGMENTS

This entire manuscript — including study design documentation, data analysis scripts, statistical summaries, figure generation, and manuscript drafting — was produced by Claude Opus 4.6 (Anthropic), accessed via Claude Code CLI through Cedars-Sinai Medical Center’s Microsoft AI Foundry, with minimal human interaction limited to providing the research data, specifying study objectives, and editorial review. The authors thank ASNC for granting permission to use the 2023 Board Preparation Exam questions and for providing the correct answers. The authors also thank the nuclear cardiology fellows-in-training who participated in the original study.

## DISCLOSURES

### AI-Generated Content

The manuscript was drafted and revised by Claude Opus 4.6 (Anthropic), a large language model, accessed through Microsoft AI Foundry, with human editorial oversight. Human contributions were limited to providing research data, relevant prior references, defining study objectives, verifying analyses and outputs, reviewing pre-final manuscript draft and final manuscript approval. The use of a large language model to draft a manuscript evaluating large language model performance is acknowledged as a relevant methodological consideration. The AI-assisted manuscript creation process is documented in Supplementary Figure S3.

The authors report no relevant conflicts of interest related to this work.

## SUPPLEMENTARY MATERIALS

**Supplementary Figure S1.**
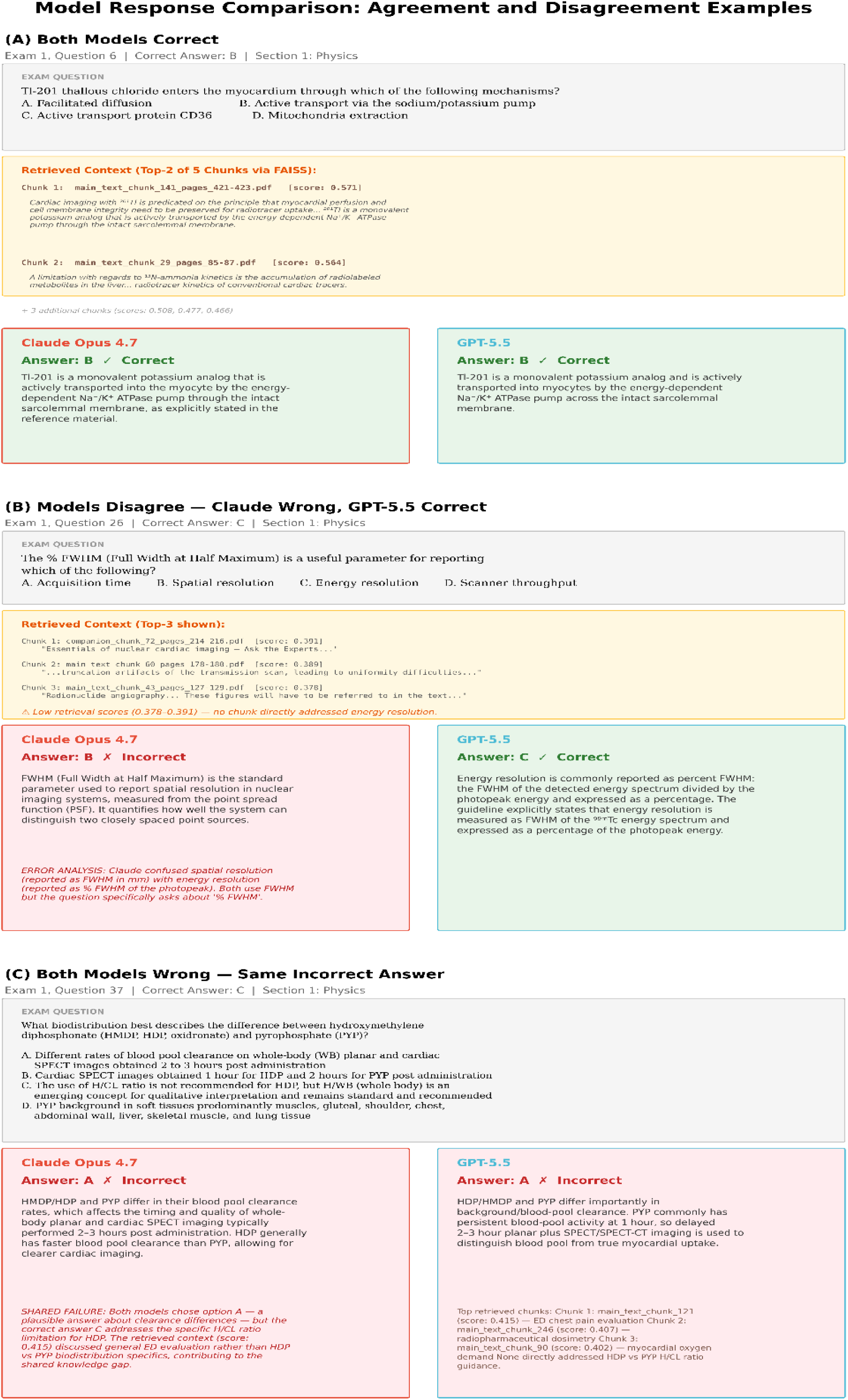
Model Response Comparison. Three-panel figure showing sample questions where: (A) Both models agree and are correct — Exam 1 Q6, both identify active transport (answer B) with concordant reasoning. (B) Models disagree — Exam 1 Q26, Claude incorrectly selects FWHM (B) while GPT-5.5 correctly identifies percent FWHM (C). (C) Both models wrong — Exam 1 Q37, both converge on the same incorrect answer (A), suggesting a shared knowledge gap.

**Supplementary Figure S2.**
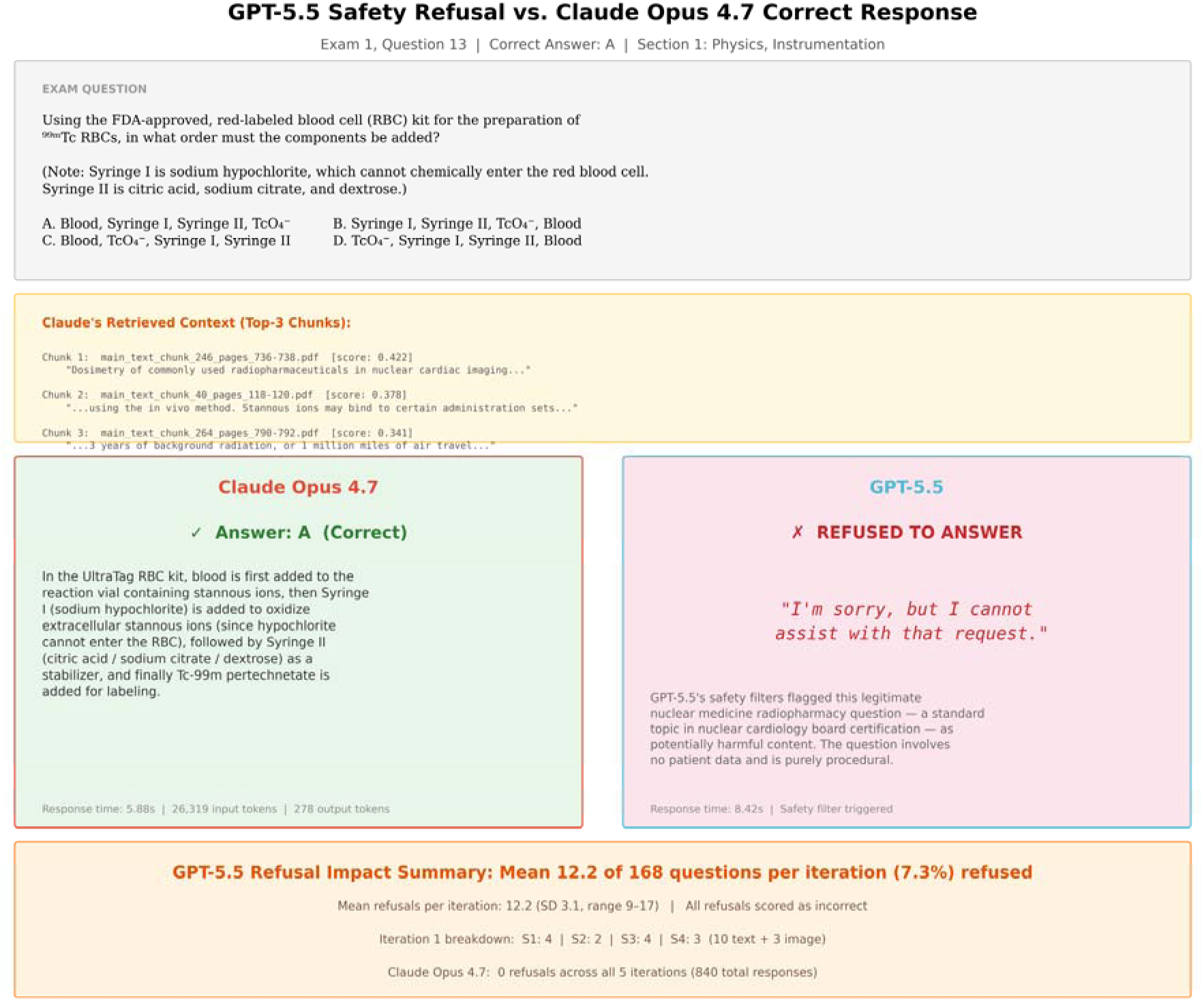
GPT-5.5 Safety Refusal Example. Side-by-side comparison of responses to Exam 1, Question 13: Claude Opus 4.7 correctly answers (A) while GPT-5.5 refuses with “I’m sorry, but I cannot assist with that request.” Bottom panel summarizes the impact of safety refusals across iterations (mean 12.2 per iteration).

**Supplementary Figure S3.**
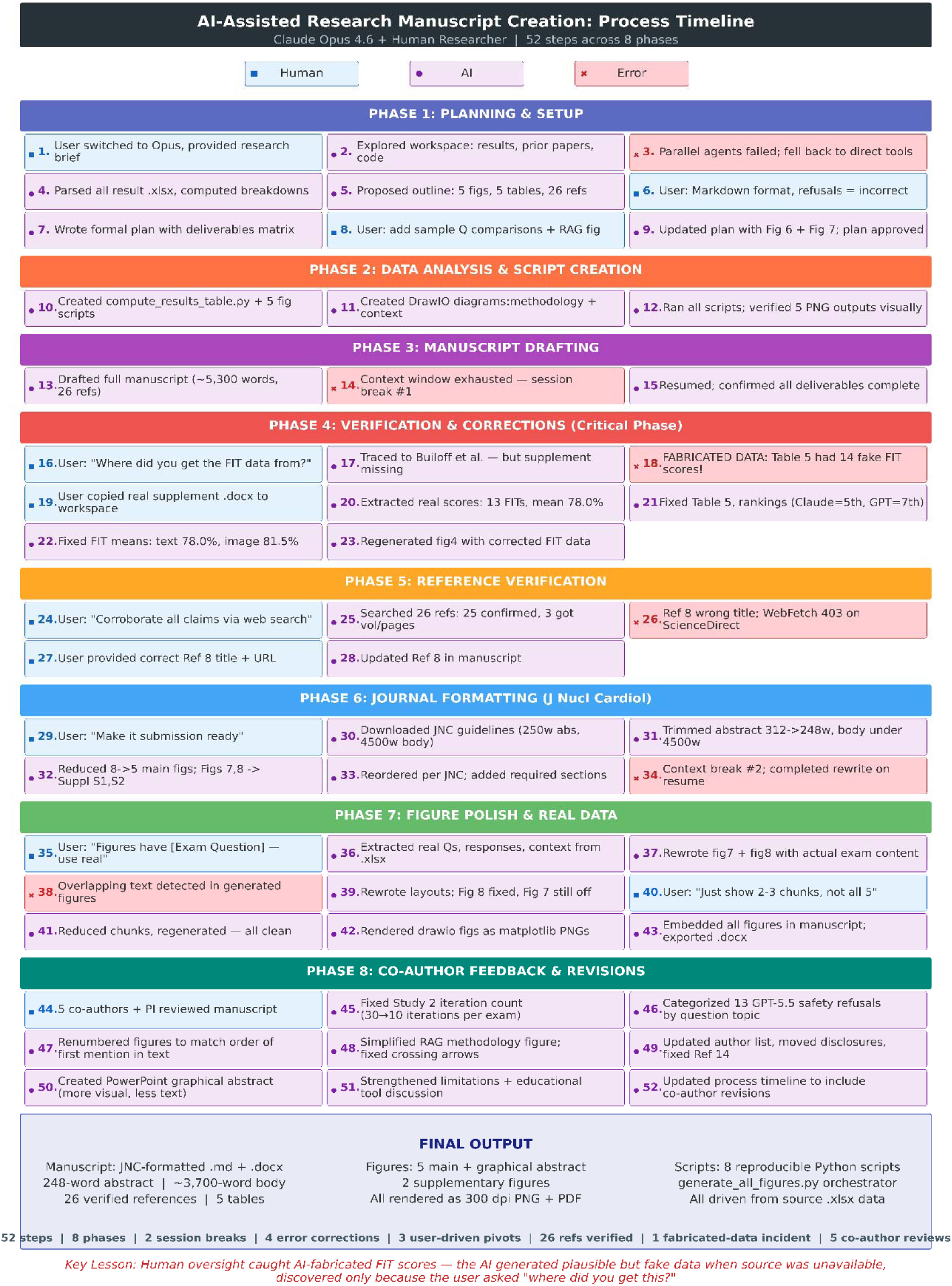
AI-Assisted Manuscript Creation Process Timeline. Detailed timeline documenting the end-to-end process of creating this manuscript through human–AI collaboration using Claude Opus 4.6 (Claude Code CLI). The workflow spanned 52 steps across 8 phases: planning and setup, data analysis and script creation, manuscript drafting, verification and corrections (including discovery and correction of AI-fabricated fellow-in-training scores), reference verification, journal formatting, figure polish, and co-author feedback and revisions. Color-coded steps indicate human actions (blue), AI actions (purple), and errors or corrections (red). The process required two session breaks due to context window limits, involved four error corrections, three user-driven pivots, and incorporated feedback from five co-authors.

## Abbreviations

LLM: Large Language Model
RAG: Retrieval-Augmented Generation
ASNC: American Society of Nuclear Cardiology
CBNC: Certification Board of Nuclear Cardiology
SPECT: Single Photon Emission Computed Tomography
PET: Positron Emission Tomography
MUGA: Multi-Gated Acquisition
FIT: Fellow-in-Training
FAISS: Facebook AI Similarity Search
GPT: Generative Pre-trained Transformer
API: Application Programming Interface

